# Contrasting the brain imaging features of MOG-Antibody disease, with AQP4- Antibody NMOSD and Multiple Sclerosis

**DOI:** 10.1101/2020.09.22.20198978

**Authors:** Silvia Messina, Romina Mariano, Adriana Roca-Fernandez, Ana Cavey, Maciej Jurynczyk, Maria Isabel Leite, Massimiliano Calabrese, Mark Jenkinson, Jacqueline Palace

## Abstract

Neuromyelitis optica associated with aquaporin-4-antibodies (NMOSD-AQP4) and myelin oligodentrocyte-glycoprotein antibody-associated disorder (MOGAD) have been recently recognised as different from multiple sclerosis.

Although conventional MRI may help distinguish multiple sclerosis from antibody-mediated diseases, the use of quantitative and non-conventional imaging may give more pathological information and explain the clinical differences.

We compared, using non-conventional imaging, brain MRI findings in 75 subjects in remission with NMOSD-AQP4, MOGAD, multiple sclerosis or healthy controls (HC). Volumetrics, white matter and cortical lesions, and tissue integrity measures using diffusion imaging, were analysed in the four groups along with their association with disability (expanded disability status scale [EDSS] and visual acuity).

The volumetric analysis showed that, deep grey matter volumes were significantly lower in multiple sclerosis (p=0.0001) and MOGAD (p=0.02), compared to HC. Relapsing MOGAD had lower white matter, pallidus and hippocampus volumes than in monophasic (p<0.05).

Optic chiasm volume was reduced only in NMOSD-AQP4 who had at least one episode of optic neuritis (ON) (NMOSD-AQP4-ON vs NMOSD-AQP4 p<0.001, HC p<0.001, MOGAD-ON p=0.04, multiple sclerosis-ON p=0.02) likely reflecting the recognised posterior location of NMOSD-AQP4-ON and its severity.

Lesion volume was greatest in multiple sclerosis followed by MOGAD and in these two diseases, the lesion volume correlated with disease duration (multiple sclerosis R=0.46, p=0.05, MOGAD R=0.81, p<0.001), cortical thickness (multiple sclerosis R=-0.64, p=0.0042, MOGAD=-0.71, p=0.005) and deep grey matter volumes (multiple sclerosis R=-0.65, p=0.0034, MOGAD R=-0.93, p<0.001).

Lesional-fractional anisotropy (FA) was reduced and mean diffusivity increased in all patients, but overall, FA was only reduced in the non-lesional tissue in multiple sclerosis (p=0.01), although focal reductions were noted in NMOSD-AQP4, reflecting mainly optic nerve and corticospinal tract pathways.

Cortical/juxtacortical lesions were seen in a minority of MOGAD, while cortical/juxtacortical and purely cortical lesions were identified in the majority of multiple sclerosis and in none of the NMOSD-AQP4.

Non-lesional FA in NMOSD-AQP4, lower white-matter volume and female sex in multiple sclerosis, and lower brainstem volume in MOGAD were the best predictors of EDSS disability (accounting for 46%, 49% and 19% respectively). Worse visual acuity associated with lower optic chiasm volume in NMOSD-AQP4 and lower thalamus volume in MOGAD (accounting for 58% and 35% respectively).

Although MOGAD patients had good outcomes, deep grey matter atrophy was present. In contrast, NMOSD-AQP4 patients showed a relative sparing of deep grey matter volumes, despite greater residual disability as compared with MOGAD patients. NMOSD-AQP4 but not MOGAD patients showed reduced FA in non-lesional tissue.

## Introduction

In the past fifteen years two new antibody mediated Central Nervous System (CNS) diseases, which had previously been thought to be multiple sclerosis variants, have been identified. The first, aquaporin-4-antibody (AQP4-Ab) disease (Lennon et al 2005), is a primary astrocytopathy and is recognised to be the major cause of the neuromyelitis optica spectrum disorders (NMOSD) (Weinshenker et al 2006, Wingerchuk et al 2015). The more recently discovered myelin oligodentrocyte glycoprotein antibody-associated disorder (MOGAD), targeting myelin, is associated with a wider clinical phenotype including NMOSD and anatomically limited forms, such as optic neuritis and transverse myelitis, acute disseminated encephalomyelitis (ADEM) and a less common isolated cortical syndrome (Jurynczyk et al 2017b, Ogawa et al 2017).

Although the symptoms across these antibody diseases and multiple sclerosis can overlap, there are important clinical differences that suggest pathological distinction. In particular, and in contrast to multiple sclerosis, the antibody-mediated diseases lack a progressive phase thus, disability is solely relapse related. Additionally, the majority of disease modifying treatment for multiple sclerosis, mainly acting on T cells, have been reported to be deleterious in NMOSD associated with AQP4-Ab (NMOSD-AQP4) (Kira 2017, Palace et al 2010), and potentially ineffective or harmful in MOGAD (Chen et al 2020, Kaneko et al 2018).

MRI plays a fundamental role in the diagnosis and the monitoring of inflammatory CNS diseases. Brain lesions have been reported in up to 60% of NMOSD-AQP4 patients (Pittock et al 2006). Characteristically, lesions localise to areas rich in AQP4 water-channel (e.g. periependymal regions, area postrema) although non-specific white matter lesions are more common (Matthews et al 2013) and a wide range of other lesions and enhancement are described (Wingerchuk et al 2015). MOGAD brain lesions can be difficult to distinguish from NMOSD-AQP4 (Jurynczyk et al 2017a) but may also appear ADEM-like (Hoftberger et al 2015). The presence of lateral and inferior temporal lobe lesions, or Dawson’s fingers or U/S shaped juxtacortical lesions, may distinguish multiple sclerosis from these antibody disorders (Jurynczyk et al 2017a). Additionally, longitudinally extensive spinal cord lesions characteristic of NMOSD-AQP4 and MOGAD are exceptional in multiple sclerosis (Asnafi et al 2020).

Although conventional MRI may identify characteristic lesions in NMOSD-AQP4 disease (Kim et al 2015), quantitative and non-conventional imaging may give more pathological information. Opinion varies as to whether NMOSD-AQP4 causes normal appearing white and grey matter abnormalities and atrophy (Chanson et al 2013, Duan et al 2012, Rocca et al 2004), probably related to whether the cohorts mix antibody positive and negative together and whether the tracts associated with lesions are accounted for (Kim et al 2017, Matthews et al 2015).

Non-conventional volumetric and diffusion imaging data in MOGAD are lacking. Understanding the differences between multiple sclerosis and these auto-antibody diseases may give clues as to why multiple sclerosis causes a progressive neurodegenerative process, not modified with current therapeutics and explain the important clinical differences. Additionally, comparing three inflammatory CNS diseases can test whether surrogates for disability measures are generic or disease specific.

This study compares and contrasts non-conventional and quantitative brain imaging features in 75 subjects across these three diseases and healthy-controls. We describe, in the remission phase: volumetrics (total and regional), white matter lesion distribution, tissue integrity using diffusion tensor imaging, cortical lesions, and the association with disability (Expanded Disability Status Scale [EDSS] and visual acuity).

## Materials and Methods

### Ethics

Participants signed informed, written consent specific to this study, under ethical approval project code: REC 17/EE/0246 and the database REC 16/SC/0224. Consent was obtained according to the Declaration of Helsinki. The reporting of this research was done in conjunction with the STROBE supporting guidelines.

### Patients

Adult subjects (≥18 years) were recruited through the Oxford Neuromyelitis Optica and Multiple Sclerosis clinics between January 2018 and August 2019. Patients with only optic neuritis without any brain or spinal cord involvement were excluded. Relapsing-remitting multiple sclerosis patients were selected to cover a similar range of EDSS scores and disease activity (number of relapses) although, the recognised differences between the two antibody cohorts (e.g. NMOSD-AQP4 is more common in female with older age at onset) meant exact matching across the three disease groups was not possible.

A total of 19 NMOSD-AQP4, 20 MOGAD, 18 multiple sclerosis and 18 healthy controls (HC) were enrolled in the study (see table 1). Testing for the presence of AQP4-Ab and MOG-Ab were performed in the Autoimmune Neurology Laboratory using a cell-based assay as described by Waters et al (Waters et al 2015, Waters et al 2012). None of the multiple sclerosis patients had AQP4-Ab or MOG-Ab, and there were no dual AQP4-Ab/MOG-Ab positive patients.

**Table 1:** baseline clinical and demographic characteristics of the enrolled participants.

|  | NMOSD-AQP4 | MOGAD | MS | HC | *p*-value |
| --- | --- | --- | --- | --- | --- |
| Participants, n | 19 | 20 | 18 | 18 | NA |
| Mean age at onset ± SD | 55.6 ± 13.2 | 41.8 ± 11.0 | 44.1 ± 6.6 | 38.9 ± 13.4 | <0.001 |
| Female, % | 68.4 | 50 | 55.5 | 55.5 | 0.695 |
| Caucasian, % | 52.6 | 100 | 100 | 83 | 0.001 |
| Median disease duration, years (range) | 11 (0-24) | 2 (0-24) | 11.5 (1-24) | NA | 0.018 |
| Median number of relapse (range) | 2 (1-11) | 2 (1-11) | 3 (1-13) | NA | 0.347 |
| Median EDSS (range) | 3 (0-7) | 1.5 (0-7) | 2.0 (0-6) | NA | 0.025 |
| Mean EDSS ± SD | 3.0 ± 1.9 | 1.7 ± 1.6 | 2.8 ± 1.7 |  | 0.055 |
| Mean LogMAR VA OD ± SD | 0.5 ± 1.2 | -0.1 ± 0.6 | -0.08 ± 0.1 | NA | 0.053 |
| Mean LogMAR VA OS ± SD | 0.4 ± 1.0 | 0.1 ± 0.7 | -0.1 ± 0.1 |  | 0.085 |

**Legend:** SD=standard deviation; NA=not applicable; MS=multiple sclerosis; VA=visual acuity;

### Clinical assessment

All the participants enrolled in the study, underwent a clinical assessment on the day of the MRI scan which included: EDSS in the disease cohorts, and the assessment of the visual acuity using the Early Treatment in Diabetes Retinopathy Study (ETDRS) charts at a 1-m distance. The logarithm of the minimum angle of resolution (logMAR) was calculated. The following clinical data were collected: age, age at onset, onset phenotype, disease duration, phenotype at the time of the MRI, number of relapses.

### MRI Imaging protocol

Brain MRI were performed at 3T (Siemens Magnetom Prisma, Erlangen, Germany) at the Oxford Centre for Functional MRI of the Brain (FMRIB) using a 64-channel receive head/neck coil. All subjects underwent the same imaging protocol, including T1-weighted, fluid attenuated inversion recovery (FLAIR), proton density (PD), double inversion recovery T2- weighted and diffusion-weighted sequences. See supplementary materials for MRI sequences parameters.

### Image analysis

Imaging data were analysed using the FMRIB Software Library of tools (University of Oxford, UK) (Jenkinson et al 2012, Smith et al 2004, Woolrich et al 2009). See supplementary materials for further details.

### Volumetric analysis

#### Total Brain Volume, White matter fraction and Grey matter fraction

The T1 sequence was bias-corrected and the lesions were filled using the manually obtained FLAIR lesion mask (see below), according to (Gelineau-Morel et al 2012). The brain tissue volume, white matter and grey matter, normalised for subject head-size, were estimated with SIENAX, part of FSL (Smith et al 2004). The total brain, white matter fraction and grey matter fraction volume, in mm^3^, were divided by 1,000,000.

#### Deep grey matter volumes

The basal ganglia volume (thalamus, caudate, putamen, pallidum, amygdala and accumbens), the hippocampus and the brainstem volumes were calculated using the automated segmentation tool FIRST, part of FSL (Patenaude et al 2011). The 3D-T1 image was bias corrected and lesion-filled (Gelineau-Morel et al 2012). The bilateral structures (thalamus, caudate, putamen, pallidum, accumbens, hyppocampus and amygdala) were averaged. All the structures were then normalized by the volumetric scaling factor obtained from SIENAX (Smith et al 2002).

#### Cortical thickness, cortical and deep white/grey matter volumes

cortical reconstruction and volumetric segmentation were performed with the Freesurfer image analysis suite (http://surfer.nmr.mgh.harvard.edu)(Dale et al 1999, Fischl et al 1999) which include a volumetric and a surface-based stream. Cortical thickness, cortical volume, the deep white/grey matter structures, 3^rd^ and 4^th^ ventricles volumes were obtained. The volumetric measures were normalised per head size using the volumetric scaling factor obtained from SIENAX. The cortical thickness was calculated for the right and the left hemisphere and then was averaged.

### Lesions analysis

#### White matter lesion analysis

FLAIR lesion masks were transformed to the MNI space, via T1 image. Number of lesions and lesion volume for each subject were obtained from the lesion mask, in MNI space. The lesion masks for all subjects were then merged and averaged to create a lesion probability map for each disease group. Three subtraction maps were obtained for the three diseases group (NMOSD-AQP4-MOGAD; NMOSD-AQP4-multiple sclerosis, MOGAD-multiple sclerosis). A voxel-wise analysis between disease groups was performed using the non-parametric permutation testing *randomise* (part of FSL) with Threshold-Free Cluster Enhancement (5000 permutations of every of the four contrast)(Winkler et al 2014). The clusters with a p-value <0.05 will be shown in the results figure 4.

#### Cortical lesions analysis

cortical lesions were counted visually on each double inversion recovery (DIR) sequence with simultaneous reference to FLAIR images and T1 images by a researcher with clinical experience in multiple sclerosis (SM) and a researcher with extensive experience and expertise in detection of cortical lesions (MC), blinded to the participant status. In case of disagreement a final consensus was reached between the raters. After scoring had been finished, scans were decoded.

### Diffusion-weighted imaging analysis: white matter and grey matter

#### Pre-processing

for diffusion pre-processing see supplementary material.

#### White matter lesions and normal white matter diffusion measures

fractional anisotropy (FA), mean diffusivity (MD), axial diffusivity (AD) and Radial diffusivity (RD) maps were created from the FMRIB’s Diffusion Toolbox (FDT) output and then registered to T1 structural image space. FA, MD, AD and RD values were then calculated within ROIs defined by the lesion mask and non-lesioned white matter tracts, as obtained from the JHU atlas.

#### Cortical and deep grey matter MD

the cortical mask was created from the right and left cortex labels, output by FreeSurfer. The deep grey matter volumes templates were created from the FIRST output. The cortex and the deep grey matter template were binarized to obtain masks. MD was calculated within the cortex and deep grey matter ROIs.

#### Tract Based Spatial Statistics

voxelwise statistical analysis of FA data was carried out using TBSS (Tract-Based Spatial Statistics)(Smith et al 2006), part of FSL. Data were compared according to group, using non-parametric permutation testing implemented by *randomise* (part of FSL) with Threshold-Free Cluster Enhancement. Lesion masks were entered as a nuisance regressor in the final model so that lesioned voxels would be disregarded in the statistical comparison of groups.

### Statistical Analysis

The statistical analysis of non-imaging data was performed using RStudio version 1.1.447. Data cleaning was performed before the data analysis, considering both range and consistency checks. Quantitative variables were described using means and SDs, median and range. Differences between means and differences between proportions were evaluated by t-test and ?2 test, respectively. A Shapiro-Wilk test was performed to assess the normal distribution of data. In case the distribution was not normal, a Wilcoxon test was performed. Linear models with sex, age as independent variables (and volumetric scaling factor in case of volumetric measures), were fit to compare the MRI findings between the four study groups. A Bartlett test was performed to assess the variance before including the variables in the linear regression model. In case of non-equal variance, the variables were transformed. The estimated marginal means were calculated based on the final model and a Bonferroni pairwise-comparison between groups was calculated. The results were expressed as estimates and standard errors. The significant *post hoc* comparisons are shown in the results section. Pearson’s correlation was used to test the relationship between MRI findings and clinical and demographic variables. For the cortical lesions’ analysis Cohen’s kappa was calculated to assess inter-rater agreement. A k<0 indicates poor agreement, 0.01–0.20 slight, 0.21– 0.40 fair, 0.41–0.60 moderate, 0.61– 0.80 substantial and 0.81–1.0 almost perfect agreement.

Finally, for EDSS, univariate linear regression models were fitted in every disease group, using MRI findings as independent variables, and EDSS as the dependent variables (see the results section for the list of demographic and MRI variables). In addition, a multivariate linear regression model with a stepwise variable selection based on the Akaike information criterion (AIC), was fitted using age and sex, and the MRI variables with p-value <0.2 in the univariate model.

For visual acuity (mean LogMAR), we use the same demographic variables and only two *a priori* MRI measures as independent variables (see the results section for the list of demographic and MRI variables) in the multivariate linear regression model.

For all analyses, p < 0.05 was considered statistically significant. Graphs were created using ggplot and ggpubr packages, Rstudio.

### Data Availability

All data is collected and stored in accordance with GDPR guidelines. Data availability is dependent on specific collaboration. Analysis software and methods are publicly available.

### Analysis plan

See study flowchart (Supplementary figure 2).

**Figure 1:**
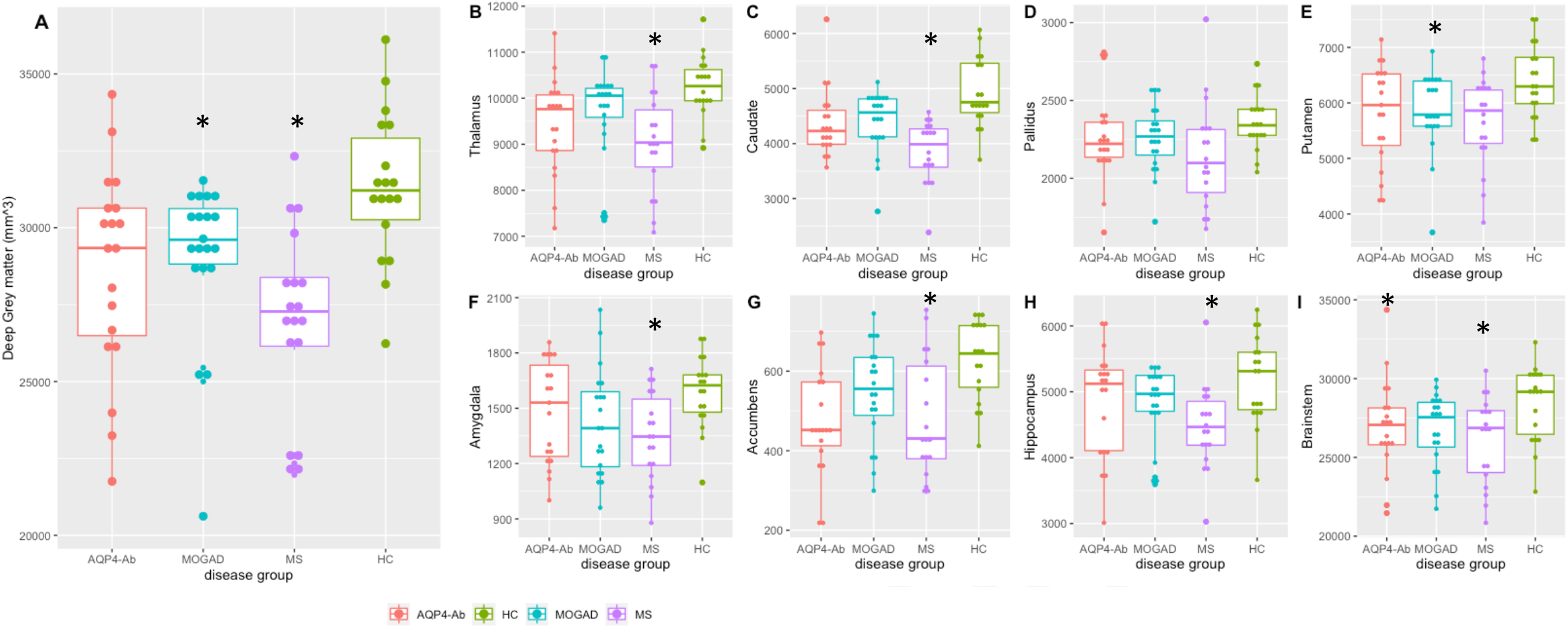
Total deep grey matter volumes (A), basal ganglia, hippocampus and brainstem volumes (B-I) between disease groups. The graphs represent the median and, the 25^th^ and 75^th^ percentile; **Legend:** * = p<0.05 when compared to HC, on the linear model adjusted for age, sex and volume scaling factors; AQP4-Ab=NMOSD-AQP4, MS=multiple sclerosis

**Figure 2:**
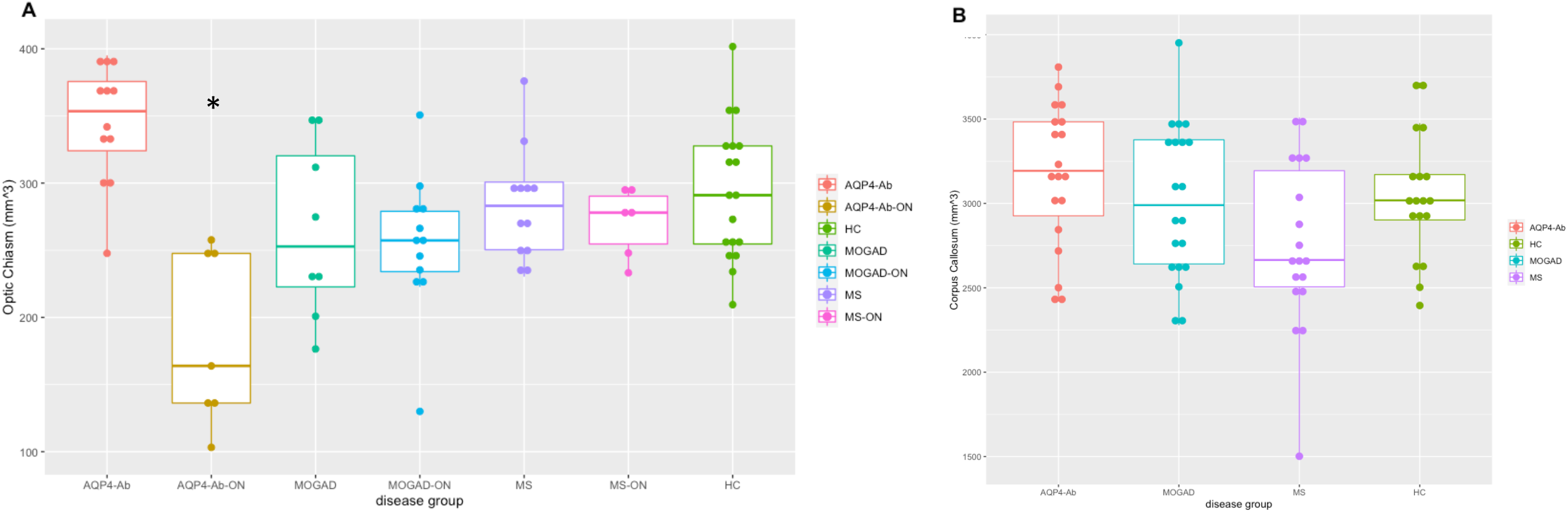
Optic chiasm and corpus callosum volumes between disease groups. The graphs represent the median and, the 25^th^ and 75^th^ percentile. In figure A optic chiasm (mm^3^) and in figure B corpus callosum (mm^3^). **Legend**: * = p<0.05 when compared to HC, MOGAD-ON and MS-ON on linear model, adjusting for age, sex and volume scaling factor. AQP4-Ab=NMOSD-AQP4, MS=multiple sclerosis

## Results

The clinical and demographic characteristics of the four group are represented in Table 1 and most features are matched across the groups. Exact matching was not possible due to the expected demographic differences between the three disease groups and the rarity of the antibody diseases. Age was not directly comparable across the groups, thus, was included in the regression model, together with sex. MOGAD is a recently recognised condition, thus the disease duration was shorter than the other groups.

### Volumetric analysis

Non-normalised volumes are shown in supplementary table 1. All group comparisons of normalised volumes used linear regression with Bonferroni correction and were adjusted for age, sex and volumetric scaling factor.

#### Total Brain Volume, white matter fraction, grey matter fraction and ventricular volumes

Supplementary figure 3 (A-C) shows a trend-level decrease in normalised brain volume (NBV), total white matter fraction (WMF) and total grey matter fraction (GMF) in NMOSD- AQP4 and multiple sclerosis patients, and no statistically significant difference in the third and fourth ventricle volumes (supplementary figure 3 D-E).

**Figure 3:**
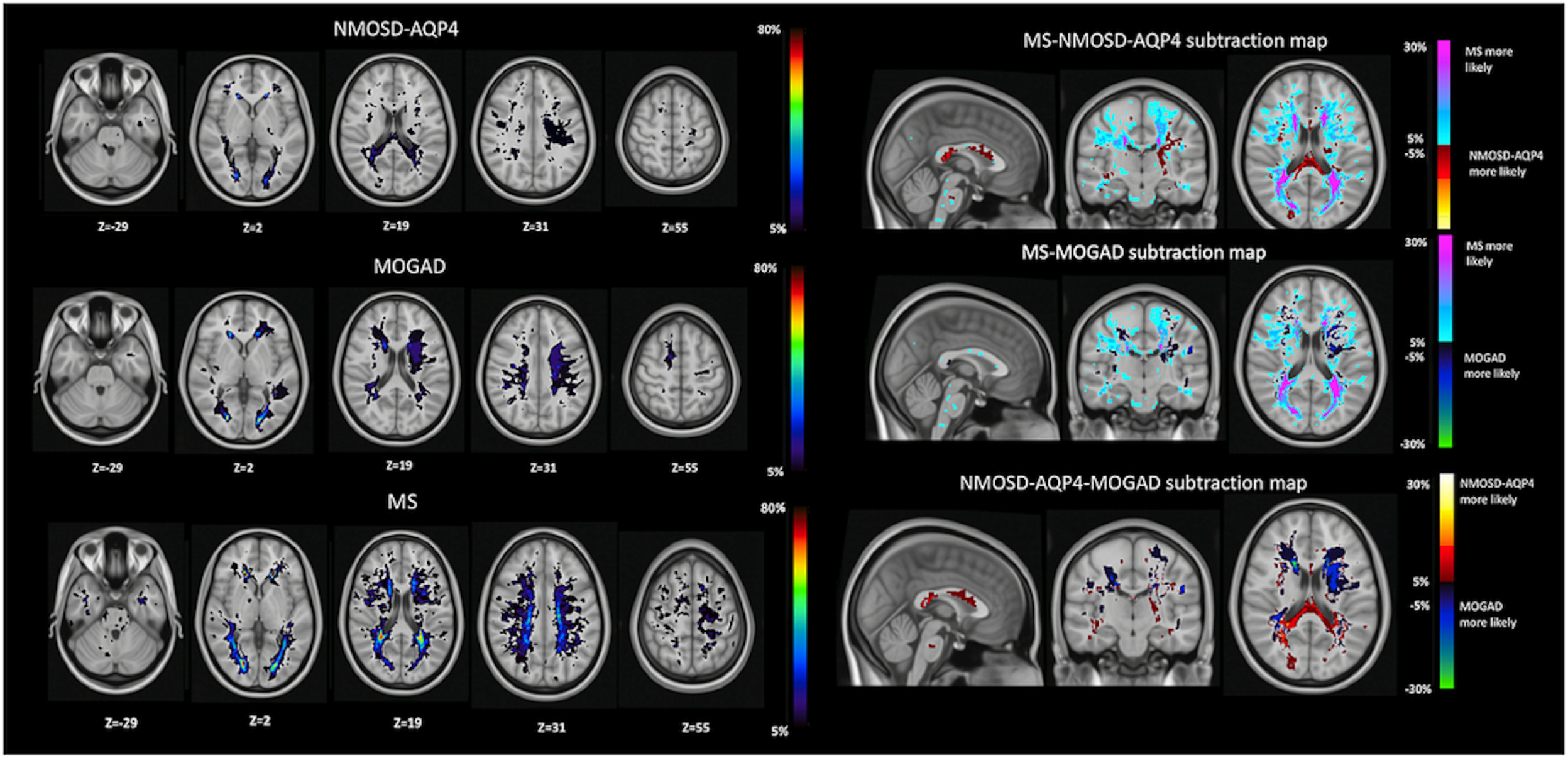
Lesion probability map in NMOSD-AQP4, MS and MOGAD; subtraction maps: MS-NMOSD-AQP4, MS-MOGAD, NMOSD-AQP4-MOGAD. **Legend**: lesions identified on the lesions map using the Talairach Atlas. MS=multiple sclerosis.

**Figure 4:**
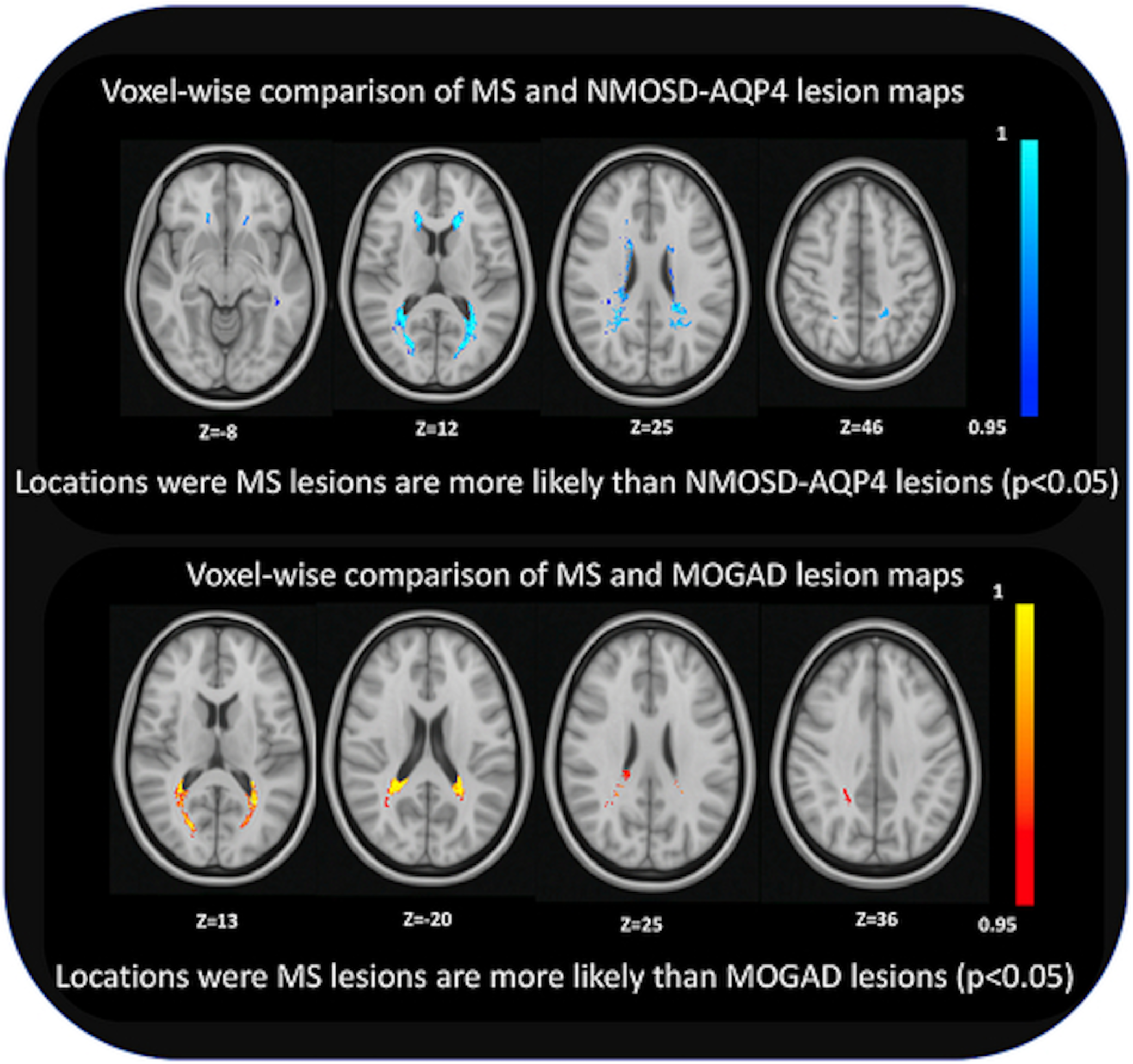
Voxel-wise comparison between NMOSD-AQP4 and MS and, MOGAD and MS. NMOSD-AQP4 and MOGAD comparison non-significant. **Legend:** MS=multiple sclerosis.

#### Grey matter volumes

when considering the total deep GM volumes, only multiple sclerosis and MOGAD showed reduced volumes when compared to HC (multiple sclerosis p=0.0001, MOGAD p=0.02) (figure 1A).

The basal ganglia, hippocampus and brainstem volumes across groups are shown in figure 1 B-I.

For the individual deep grey matter structures, compared to HC, thalamus, caudate, hippocampus, accumbens, amygdala and brainstem were all significantly smaller in multiple sclerosis (p=0.0008, p<0.001, p=0.007, p=0.01, p=0.007 and p=0.01 respectively), while the putamen was only smaller in MOGAD (p=0.050) and the brainstem was only smaller in NMOSD-AQP4 (p=0.04). Moreover, the putamen showed a trend for smaller values in multiple sclerosis (p=0.06), as did the thalamus and amygdala with smaller values in NMOSD- AQP4 (p=0.09 and p=0.07), and the caudate, amygdala and brainstem with a trend for smaller values in MOGAD (p=0.07, p=0.08 and p=0.08). Compared to multiple sclerosis, the caudate is significantly smaller in NMOSD-AQP4 (p=0.01) and showed a trend for smaller values in MOGAD (p=0.07), and there were no significant differences between NMOSD-AQP4 and MOGAD.

The analysis of the deep grey matter structures, repeated with non-normalised data and including sex, age and volume scaling factors, yielded similar results.

#### Cortical thickness and volumes

cortical thickness was not different between groups, however cortical volume was smaller in NMOSD-AQP4 when compared to the other groups at the trend level (p=0.08) (see supplementary figure 4 A-B).

#### White matter region of interest volumes

given that visual function is often involved in the antibody diseases, and association fibres can be abnormal in multiple sclerosis, we selected the optic chiasm and corpus callosum as regions of interest for volume analysis.

No significant difference in optic chiasm volumes were seen. When only including patients with at least one episode of optic neuritis (NMOSD-AQP4-ON, MOGAD-ON and multiple sclerosis-ON) NMOSD-AQP4-ON had significantly lower chiasmatic volumes when compared to the other groups (NMOSD-AQP4 vs NMOSD-AQP4-ON p<0.001, NMOSD- AQP4-ON vs HC p<0.001, NMOSD-AQP4-ON vs MOGAD-ON p=0.04, NMOSD-AQP4-ON vs multiple sclerosis-ON p=0.02) likely reflecting the recognised posterior location of NMOSD-AQP4-ON and its severity (see figure 2A and table 1). We noted a reduction in corpus callosal volumes in multiple sclerosis compared to NMOSD-AQP4 (multiple sclerosis vs NMOSD-AQP4 p=0.06, multiple sclerosis vs HC p=0.2) (see figure 2B)

### Lesion analysis

#### Lesion volume

all multiple sclerosis and NMOSD-AQP4, and 14 out of 20 (70%) of MOGAD patients had brain lesions at the time of the scan. Out of the six MOGAD patients who had normal brain appearance at the time of the scan, three had brain lesions during the acute phase. The multiple sclerosis patients had a higher lesion volume when compared to NMOSD-AQP4 disease (p=0.03) and MOGAD patients (p=0.0005). Lesion volume was positively correlated with disease duration in multiple sclerosis (Pearson’s correlation R=0.46, p=0.05) and MOGAD patients (Pearson’s correlation R=0.81, p<0.001), while it showed a moderate correlation with the number of relapses in NMOSD-AQP4 (Pearson’s correlation R=0.53, p=0.02) and multiple sclerosis (Pearson’s correlation R=0.48, p=0.059).

#### Lesion probability maps

In the NMOSD-AQP4 group the lesion map showed a widespread distribution, with the highest percentage of patients having lesions in the lateral ventricle anterior horns (31.5%), and in the right lingual gyrus of the occipital lobe (42%) and 10.5% in the corpus callosum.

In the MOGAD group the lesion map showed a widespread distribution, with the highest percentage of patients having lesions in the lingual gyrus bilaterally (50% right and 57.1% left), 42.8% have small lesions in the anterior horns of the right and left lateral ventricles, 14.3% in the superior frontal gyrus and 14.3 % in the medial frontal gyrus.

In the multiple sclerosis group the lesion map showed a more focal distribution, with the highest percentage of patients having lesions in the periventricular anterior horns area (88.8% on the right and 94.4% on the left lateral ventricles), 16.6% in the brainstem, 11.1% cerebellum, and 55.5% in the corpus callosum (see figure 3)

Subtraction maps (figure 3) were created subtracting the averaged NMOSD-AQP4 and MOGAD map from the multiple sclerosis map and the averaged NMOSD-AQP4 from the averaged MOGAD map. Compared to the other two disease groups, the lesions in the NMOSD- AQP4 group are more likely to occur in the periependymal area of the corpus callosum. In multiple sclerosis, compared to both groups, lesions are more likely to occur in the periventricular and juxtacortical areas.

The voxel-wise analysis between NMOSD-AQP4 and MOGAD was not significant, while multiple sclerosis analysis showed that lesions are statistically more frequent in the periventricular areas than NMOSD-AQP4 and MOGAD (p<0.05) (see figure 4).

#### Cortical lesions analysis

a total of 69 DIR sequences were available for the cortical lesions’ analysis (16 HC, 16 NMOSD-AQP4, 19 MOGAD and 17 multiple sclerosis). See supplementary table 2 for the distribution of cortical lesions between groups. The inter-rater agreement was k=0.78 (p<0.001) with 95% confidence interval (CI) 0.59-0.97. We did not find any cortical lesions in the NMOSD-AQP4 and HC group. Three MOGAD patients (15.7%) showed only cortical/juxtacortical lesions (type I; one with one curvilinear and one with one ovoid lesion, and one participant with three curvilinear lesions), while 65% of multiple sclerosis patients had identifiable cortical/juxtacortical lesions, with three patients presenting at least one pure cortical lesion (type II) and one an extensive subpial lesion (see supplementary figure 5).

**Table 2:** summary of the main findings

|  | NMOSD-AQP4 | MOGAD | MS |
| --- | --- | --- | --- |
| Global volume measures |  |  |  |
| Brain Volume | ↓ | ↔ | ↓ |
| White Matter Volume | ↓ | ↔ | ↓ |
| Grey matter Volume | ↓ | ↔ | ↓ |
| 3<sup>rd</sup> Ventricle Volume | ↑ | ↑ | ↑ |
| 4<sup>th</sup> Ventricle Volume | ↔ | ↔ | ↔ |
| Grey matter measures |  |  |  |
| Deep grey matter Volume | ↓ | ↓↓ | ↓↓ |
| Brainstem Volume | ↓↓ | ↓↓† | ↓↓ |
| Cortex Volumes | ↓↓† | ↔ | ↔ |
| Cortex Thickness | ↓ | ↔ | ↓ |
| Cortical lesions | ↔ | ↑ | ↑↑ |
| Cortex MD | ↔ | ↔ | ↑ |
| Basal ganglia MD | ↔ | ↔ | ↔ |
| White matter measures |  |  |  |
| Optic Chiasm Volume in ON | ↓↓ | ↔ | ↔ |
| Corpus Callosum Volume | ↔ | ↔ | ↓ |
| White matter lesions Volume | ↑ | ↑ | ↑↑ |
| White matter lesion FA | ↓ | ↓ | ↓ |
| White matter lesion MD | ↑ | ↑ | ↑↑† |
| White matter lesion RD | ↑ | ↑ | ↑ |
| White matter lesion AD | ↔ | ↔ | ↔ |
| Normal appearing white matter FA | ↓ | ↔ | ↓↓ |
| Normal appearing white matter MD | ↑ | ↔ | ↑ |
| Normal appearing white matter RD | ↑ | ↔ | ↑ |
| Normal appearing white matter AD | ↔ | ↔ | ↔ |

**Legend:** compared to HC ↔ no change, ↓ reduced but not significant; ↓↓† trend, ↓↓ significantly reduced; MS=multiple sclerosis.

**Figure 5:**
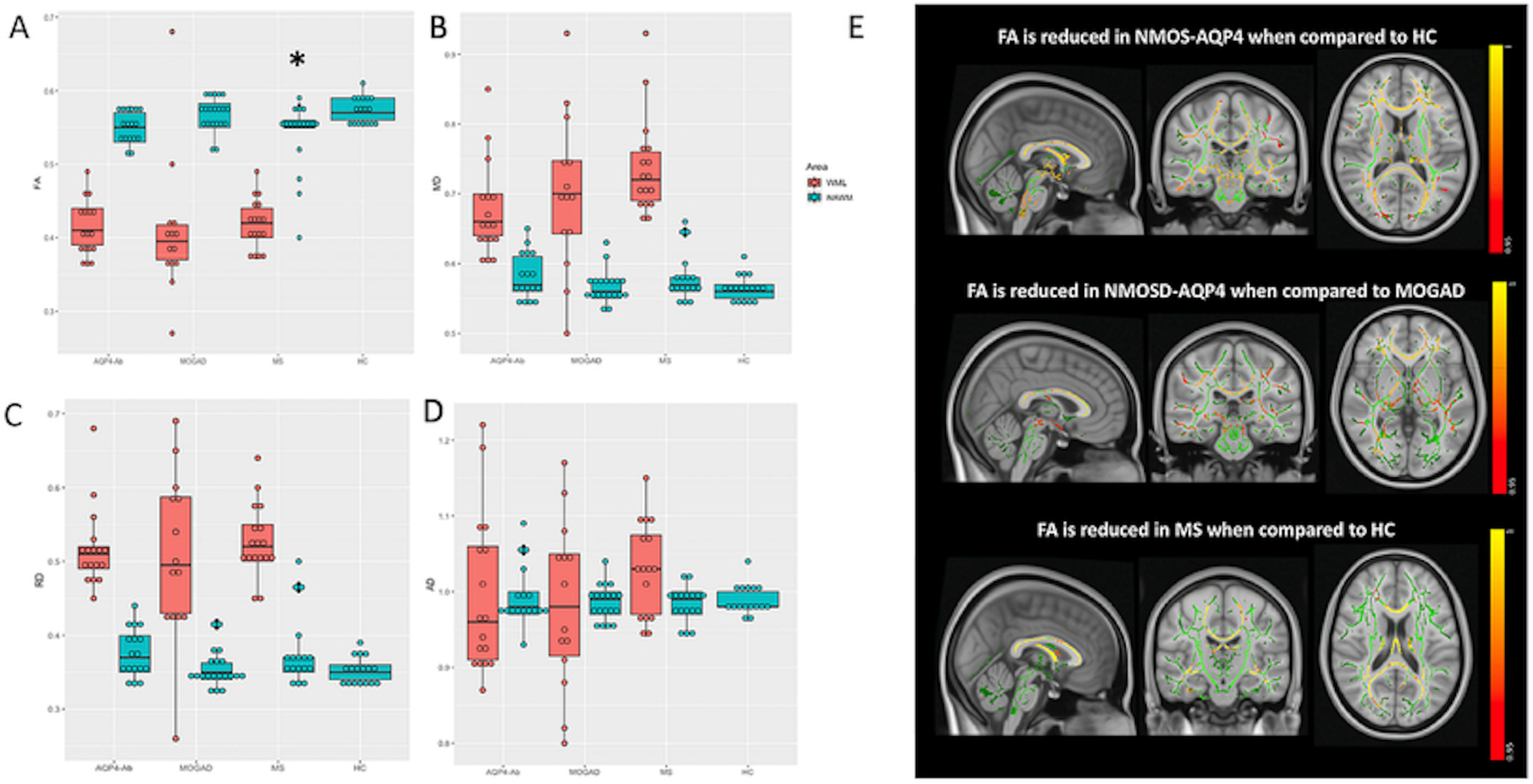
FA, MD RD and AD WML and NAWM comparison in the four groups (A-D); tract based spatial statistics FA in NMOSD-AQP4 vs HC, NMOSD-AQP4 vs MOGAD, and MS vs HC (E). **Legend:** * = p<0.05 when compared to HC, on the linear model adjusted for age and sex (A- D). The FA skeleton is shown in green with a threshold of 0.2, lower FA is shown in yellow- red with a significance of p<0.05 (E). AQP4-Ab=NMOSD-AQP4; MS=multiple sclerosis.

### Diffusion-weighted imaging analysis

All group comparisons used linear regression with Bonferroni correction and were adjusted for age and sex.

#### White matter diffusion metrics

FA and MD, AD and RD were calculated (see supplementary table 3) within the white matter lesion (WML) voxels using the FLAIR lesion masks, and also in the normal appearing white matter (NAWM).

WML FA was reduced, and WML MD and RD were increased when compared to the NAWM within all the three diseases (and also compared to HC p<0.001 in each group), but no differences were found in AD values. The WML MD in multiple sclerosis was higher than in NMOSD-AQP4 (p=0.08) at a trend level.

The multiple sclerosis NAWM-FA was lower than HC and MOGAD (p=0.01, p=0.03), while there were no differences in NAWM MD and NAWM RD, and NAWM AD (see figure 5 A-D).

#### Tract-based spatial statistics of NAWM FA

TBSS voxel-wise analysis of FA NAWM showed a significant reduction in NMOSD-AQP4 when compared with HC in the external capsule, corona radiata, the anterior limb of the internal capsule, posterior limb of the internal capsule (including fibres of the optic radiation), retrolenticular internal capsule, corpus callosum, posterior thalamic radiation (including the optic radiation), and in the brainstem, the cerebral peduncle, the middle cerebellar peduncles, medial lemniscus, the pontine crossing tracts (corticospinal tracts) and the inferior cerebellar peduncles.

FA NAWM in NMOSD-AQP4 when compared to MOGAD, showed a more widespread significant reduction (see figure 5E).

The TBSS voxel-wise analysis in the MOGAD did not show any significant difference when compared with HC.

Multiple sclerosis compared to HC, showed a significant reduction of the FA in the corpus callosum, corona radiata, and posterior thalamic radiation (see figure 5E), and a trend for a reduction in the posterior part of the corpus callosum when compared to MOGAD (p=0.07, figure not shown).

#### Grey matter diffusion metrics

MD values were analysed in the cortical and deep grey matter structures. Except for a trend-level increase in MD value in the multiple sclerosis cortex, there were no other differences, although three multiple sclerosis patients showed higher values in the deep grey matter (all three had EDSS>4) (see supplementary figure 6).

**Figure 6:**
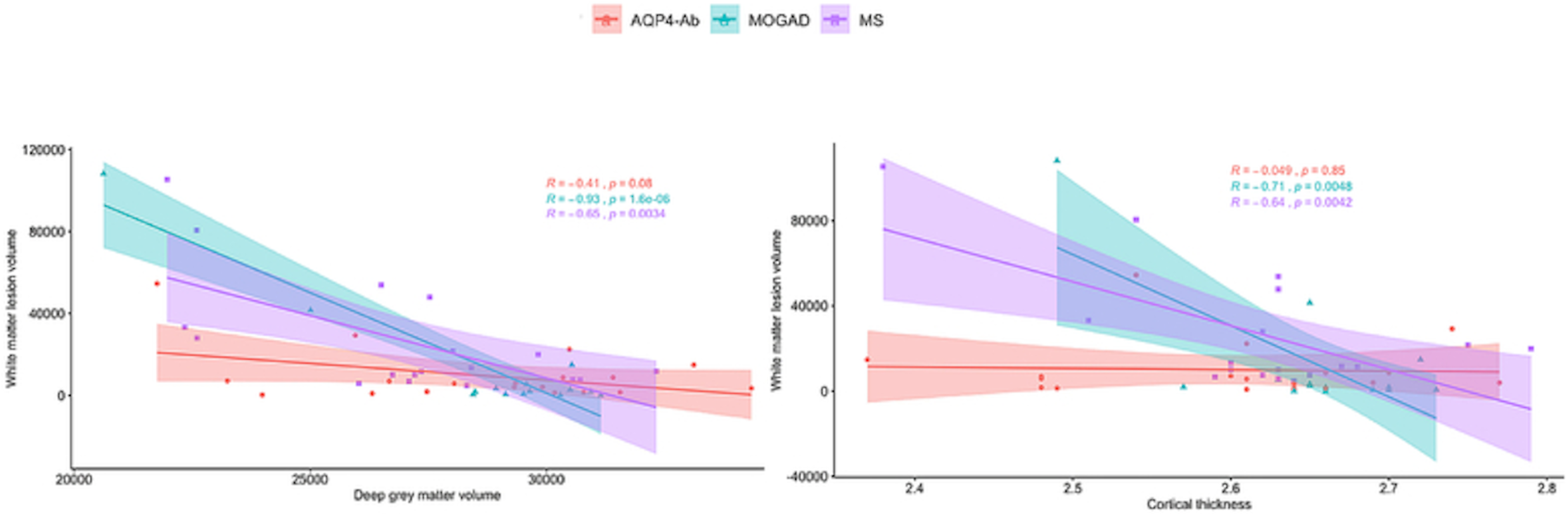
The graphs represent the correlation between lesion volume and deep grey matter volume (figure A) and between lesion volume and cortical thickness (figure B). R=Pearson ‘s correlation coefficient; The bold lines show the linear correlation for the three diseases, the areas around the lines represent the 95% confidence interval. **Legend**: AQP4-Ab=NMOSD-AQP4; MS=multiple sclerosis.

### Correlations between lesion volume and grey matter volumes in NMOSD-AQP4, MOGAD and multiple sclerosis

Because the predominant differences in non-lesional volumes were in grey matter structures we looked at the relationship between lesion volume and deep grey matter volumes and cortical volume and thickness, and found significant negative correlations in multiple sclerosis and MOGAD for deep grey matter volumes (Pearson’s correlation coefficients; multiple sclerosis R=-0.65, p=0.0034, MOGAD R=-0.93, p<0.001) and also for cortical thickness (multiple sclerosis R=-0.64, p=0.0042, MOGAD=-0.71, p=0.005) (see figure 6). No significant correlations were found with cortical volume.

### Clinical outcomes and their association with the imaging findings

#### Disability - *EDSS*

NMOSD-AQP4 had a higher EDSS (3.0 ± 1.9) compared to multiple sclerosis (2.8 ± 1.7) and MOGAD (1.7 ± 1.6) (see table 1).

We explored the association of every separate MRI finding with EDSS, within each disease group, using a univariate regression model to establish the absolute and relative strengths of the association between EDSS and each MRI variable.

The following MRI findings were used as independent variables in each univariate model: NBV, WMF, GMF, third and fourth ventricle volume, deep grey matter volumes, brainstem volume, cortical volume, cortical thickness, cortical lesions, corpus callous volume, WML volume. For diffusion measures grey matter MD was included (cortical and also deep grey matter) and to avoid collinearity for white matter we only included FA (WML FA and NAWM FA) (for the univariate analysis and the MRI findings included in each group multivariate analysis see supplementary table 4-9).

The stepwise multivariate analysis, including only variables with p=<0.2 in the univariate analysis, showed that lower NAWM FA in NMOSD-AQP4 (R-squared=0.46, p<0.001; NAWM FA slope=-61.66, p<0.001), lower WMF and female sex in multiple sclerosis (R- squared=0.49; slope=-15.69, p=0.03; beta value for sex=1.97, p=0.006) and lower brainstem volume in MOGAD (R-squared=0.19; slope= -0.0003, p=0.03), are associated with worse EDSS.

### Disability *- Visual acuity*

LogMAR VA in the NMOSD-AQP4 group was higher (worse) (VA right eye=0.46 ± 1.18, VA left eye=0.45 ± 1.03) than in MOGAD (VA right eye=-0.11 ± 0.56, VA left eye 0.05 ± 0.70) and multiple sclerosis (VA right eye=-0.08 ± 0.15, VA left eye=-0.10 ± 0.11) (Table 1). In particular, in the NMOSD-AQP4 group 6/19 have a visual acuity worse than 6/60 in at least one eye and one participant has no light perception bilaterally.

As the thalamus, is part of the visual pathway, projecting to the occipital cortex, we explored the association of thalamus volume and optic chiasm volume, within each disease group. We built a stepwise multivariate regression, including thalamus volume, optic chiasm volume, age and sex.

We found that lower optic chiasm in NMOSD-AQP4 (R-squared=0.58, p=0.0006; estimate=- 0.007, p<0.001) and, lower thalamic volume and female sex in MOGAD (R-squared = 0.35, p=0.009, thalamic volume =-0.0004, p=0.006, beta value for sex=0.48, p=0.04) are predictors of worse visual outcome. We did not find any significant predictor in multiple sclerosis (see supplementary tables 10-11).

### Monophasic and relapsing MOGAD

Because MOGAD may be monophasic (n=6 in our group) or relapsing (n=14 in our group) we analysed the brain measures in these two subsets, first comparing both groups overall, and then dividing the relapsing group into those with brain/brainstem relapses and those with non-brain/brainstem relapses. The latter compared to monophasic patients would adjust for differences simply being due to further local relapse damage and explore if monophasic MOGAD includes some patients with a different pathological disease process to those with relapsing disease.

White matter fraction (supplementary figure 7A, p=0.04), pallidus (supplementary figure 7C, p=0.04) and hippocampus (supplementary figure 7D, p=0.005) were significantly lower, while the total deep grey matter (p=0.1) and accumbens (p=0.06) showed a trend towards reduction when comparing the total relapsing group to the monophasic group.

These differences held when comparing the monophasic to the brain/brainstem relapse group (supplementary figure 7 F-J; deep grey matter, p=0.03; white matter, p=0.08; pallidus, p=0.004; hippocampus, p=0.01; accumbens, p=0.08).

When comparing the monophasic to the non-brain/brainstem relapses, there was a trend for hippocampal volumes to be lower in the non-brain/brainstem relapsing group (supplementary figure 7N, p=0.09). However, the caudate was significantly lower in the monophasic group (p=0.03) (figure not shown).

## Discussion

This is the first study using quantitative and non-conventional MRI to compare and contrast MOGAD disease, NMOSD-AQP4 and multiple sclerosis and showed different patterns of change across the three diseases (Table 3). We noted significant volume loss in the deep grey matter structures in multiple sclerosis followed by MOGAD but not NMOSD-AQP4, despite greatest disability being seen in NMOSD-AQP4 and least in MOGAD. Relapsing MOGAD had lower white matter, pallidus and hippocampus volumes than in monophasic disease and this difference held for hippocampal volumes even when only those with non-brain relapses were included. In the white matter, optic chiasm volume was reduced only in NMOSD-AQP4- ON patients. Lesion volume was greatest in multiple sclerosis followed by MOGAD and in these two diseases, the lesion volumes correlated with disease duration, with cortical thickness and deep grey matter volumes. Only NMOSD-AQP4 lesion volume correlates with relapse numbers. Fractional anisotropy was reduced and mean diffusivity increased in lesions of all patients but, overall FA was only reduced in the non-lesional tissue in multiple sclerosis, although focal reductions were noted in NMOSD-AQP4 patients, reflecting mainly optic nerve and corticospinal tract pathways. Cortical/juxtacortical (type I) fluffy, curvilinear lesions were seen in a minority of MOGAD, while cortical/juxtacortical and purely cortical (type II) ovoid lesions were identified in the majority of multiple sclerosis patients and in none of the AQP4- NMOSD patients. EDSS disability could be predicted by 46% in the NMOSD-AQP4 group using NAWM FA alone, by 49% using WMF and female sex in multiple sclerosis, and by 19% using brainstem volume in MOGAD. Visual acuity was predicted by 58% using optic chiasm volume in NMOSD-AQP4 disease and by 35% using thalamic volume and female sex in MOGAD.

Our study confirmed that grey matter volume, particularly deep grey matter structures, was most sensitive to atrophy in multiple sclerosis (Messina & Patti 2014, Zhang et al 2020) and, for the first time, this was identified as the only region of atrophy in MOGAD. Of note deep grey matter volume was lower, but not significantly reduced in NMOSD-AQP4, despite poorer outcomes, probably related to the greater group variability, but it appears this measure is less sensitive in NMOSD-AQP4 in keeping with other published papers (Duan et al 2012, Eshaghi et al 2016, Matthews et al 2015).

It is intriguing to note that grey matter volume changes occurred in the two conditions thought to primarily target myelin – a predominant white matter component. However, wider pathological CNS involvement in multiple sclerosis is well recognised while, the expression of MOG throughout the CNS needs further study. Grey matter lesions have been described in ADEM and over half of children with ADEM have MOG-antibodies (Hacohen et al 2017, Salama et al 2019b). The white matter lesions in both these conditions, but not in NMOSD- AQP4 correlated negatively with deep grey matter volume suggesting current WML could be driving the atrophy by disruption of the white matter bundles projecting into the deep grey matter structures. Of interest is that in spinal cord, grey matter atrophy is also found in MOGAD (personal communication Mariano R. et al 2020), suggesting a predilection for the grey matter structures in this condition.

Only in multiple sclerosis total NAWM was found to be significantly abnormal when using FA, a sensitive measure of demyelination, and this could represent widespread lesion related damage (related to remote lesions or ‘resolved’ lesions) and/or non-lesional pathology. The tract-based analysis did identify damage in NMOSD-AQP4 but predominantly affecting the visual and pyramidal pathways, supporting remote lesional damage in areas commonly affected in this disease. This predominance of visual and pyramidal pathway tract remote involvement is supported by other studies (Kim et al 2017, Matthews et al 2015).

FA changes were not found in MOGAD NAWM, even though MOGAD attacks are equally severe at nadir (Jurynczyk et al 2017b) and 8/20 of our patients had acute large lesions, and this is in keeping with their good clinical recovery.

Of interest is that the lesions themselves across all diseases, including MOGAD, appeared similarly affected on diffusion metrics and were abnormal when compared to the NAWM measures. This suggests that the damage from permanent lesions was similar. This was reflected in all measures, except axial diffusivity and may reflect predominant myelin rather than axonal damage in the lesions.

Cortical lesions were (as expected) identified in the majority of multiple sclerosis patients. We found none in NMOSD-AQP4. Of note we found them in three MOGAD patients. This again is intriguing, considering multiple sclerosis and MOGAD may have similar predominantly white matter targets (i.e. myelin) but cortical involvement is now well described in multiple sclerosis (Calabrese et al 2010) and cortical signal changes associated with seizure have been reported as a rarer isolated phenotype in MOGAD (Ogawa et al 2017). Additionally, cortical/juxtacortical lesions have been described in clinical MRI (Salama et al 2019a). The cortical lesions may differ between these two diseases but need further study; we noted that most of the MOGAD cortical lesions were curvilinear and fluffy, involving adjacent juxtacortical regions (type I) whereas in multiple sclerosis they were mainly ovoid, and some were purely cortical (type II). Cortical/juxtacortical lesions have been described as absent in NMOSD (Calabrese et al 2012) or have been identified in only a minority of NMOSD-AQP4 patients during the acute phase (Kim et al 2016). Although, cortex is rich in AQP4 water-channel, we did not find lesions, but we reported lower cortical volume. It is possible, that the disruption of water homeostasis, due to the presence of antibodies directed against the cortex water-channel, is the key pathological mechanism in NMOSD-AQP4 cortex, and may explain the reduction of volume in NMOSD-AQP4 patients, in the absence of a disruptive lesion damage (Popescu et al 2010).

The lesion probability subtraction maps showed that lesions are more likely to be present in the periependymal area of the corpus callosum in NMOSD-AQP4 where AQP4 is highly expressed. Multiple sclerosis showed a high number of lesions in the periventricular area in line with previous findings when compared to NMOSD-AQP4 (Matthews et al 2013) and also when compared to MOGAD. We did not show a specific lesion pattern distribution in in MOGAD, and this is in part in line, with previous findings where a disperse lesions distribution was identified. Although infratentorial lesions are common in the acute phase, we did not find it in our MOGAD cohort as, in contrast to previous retrospective studies (Yang et al 2020), we only included patients in the remission phase. The location of lesions outside of the acute phase, i.e. persistent lesions, is likely to be more useful in associating them with long-term pathological effects on disability.

White matter regions of interest volumetric analysis focused on the optic chiasm and corpus callosum, being predominant brain areas involved in NMOSD and multiple sclerosis respectively and only showed a significant reduction in NMOSD-AQP4 patients who have had an ON attack. This is in keeping with the clinical residual severity of ON associated with AQP4 antibodies and the lack of abnormality outside of ON attacks, consistent with observed lack of silent damage outside of relapses in NMOSD. The observed optic radiation damage may therefore being a consequence of the retrograde degeneration, due to the involvement of the axonal loss in the optic chiasm (Juenger et al 2020).

The lack of abnormality in the optic chiasm in multiple sclerosis and MOGAD patients having ON reflects their better recovery and the lack of posterior involvement typically seen with NMOSD-AQP4 (Khanna et al 2012).

MOGAD can present with a monophasic or relapsing phenotype. Relapses can occur in up to 50% (Cobo-Calvo et al 2018, Jurynczyk et al 2017b) of patients and the risk is higher within the first year since the onset attack. Those who become antibody negative are less likely to relapse (Jurynczyk et al 2017b, Mariotto et al 2017). We showed that, relapsing patients have lower brain volumetric measures. Additionally, when removing those with relapses that involved either the brain or brainstem, relapsing disease still associated with lower hippocampal volumes and, the lack of significance maybe related to small numbers within these subgroups. Longitudinal studies may determine if hippocampal atrophy after a single attack is a predictor of relapsing disease and may help identify who should be immunosuppressed from onset.

The different MRI predictors of disability within the individual disease groups, suggests different pathological mechanism might be contributing to the tissue damage across these diseases. It is not surprising that NAWM FA was associated with EDSS in NMOSD-AQP4 disease, because the regional changes were identified in those tracts mainly associated with the optic nerve and corticospinal tract and might relate to remote damage from attacks of ON and transverse myelitis. The predictive value was high at 46%.

WMF and female sex, contributed to 49% of the EDSS in multiple sclerosis, possibly suggesting a different pathological explanation for disability when compared to NMOSD- AQP4.

Brainstem volume was the only MRI measure associated with EDSS in MOGAD, and this may reflect its eloquent site for ambulatory disability. This is also in line with our observation that the presence of brainstem lesions is associated with a worse recovery from transverse myelitis attacks (Mariano et al 2019).

In the NMOSD-AQP4 group lower optic chiasm volumes can explain 58% of worse visual acuity, in line with previous observation of a preferential involvement of the chiasm in NMOSD-AQP4 (Khanna et al 2012).

Interestingly, only lower thalamic volume was associated with MOGAD. Although, only 35% of the visual disability may be explained by the model, this intriguing finding may suggest a possible involvement of axon from the lateral retina, which remain ipsilateral and do not cross the chiasm (Kupfer et al 1967).

The main limitation of our study is the small sample size per disease group related to our selecting four sub-groups, two of which are rare diseases, made even rarer by enriching the antibody disease cohorts with patients with brain lesions when many have sparing of the brain. The use of four different groups however, allowed comparison of three inflammatory diseases against healthy controls and each other; one likely T-cell driven against myelin and two auto- antibody conditions, one against myelin and one against astrocyte antigens. This gives interesting insights into pathological differences, reflected in non-conventional and quantitative imaging measures. Although relatively small numbers, the homogeneity of a single centre study using the same scanner and protocol, along with reliable diagnostic ability (using the same expert clinicians and highly accurate assays) is an advantage and will reduce ‘noise’ and improve power. Observations from our study can then be used to build hypotheses and design future multicentre studies. Additionally, it was not possible to match all characteristics across the groups, i.e. NMOSD-AQP4 disease affects older patients and has a greater preponderance of females and is more severe, but we were able to account for this by adjusting for age and sex in the analyses.

## Conclusions

MOGAD disease is clinically less severe than NMOSD-AQP4, often with patients recovering completely, and the conventional MRI showing resolution of the brain and spinal cord lesions. Our study shows for the first time in MOGAD that deep grey matter atrophy can occur and that only lesional and not NAWM tissue damage can be detected using FA. It also highlights the relative sparing of deep grey matter in NMOSD-AQP4 disease, despite being associated with greater disability and notes focal NAWM white matter changes. Future studies should focus on the association of “invisible” symptoms (i.e. cognitive impairment and fatigue) with deep grey matter changes in MOGAD. Additionally, larger imaging studies in MOGAD and NMOSD-AQP4 patients may identify differences across the different clinical phenotype subgroups and identify new markers of damage that would be useful in clinical trials.

## Acknowledgements

we gratefully acknowledge all patients, relatives and healthy volunteers who participated in this study. We thank the radiographers Michael Sanders, Jon Campbell, David Parker and all the staff at the Wellcome Centre for Integrative Neuroimaging. We thank Dr Ludovica Griffanti for her insight on the MRI analysis, Dr Michael Cottar and Dr Hanna Nowicka for their support on diffusion analysis and Mr Fidel Alfaro Almagro for his insight on the FreeSurfer software.

---

|  | Item No | Recommendation | Page No |
| --- | --- | --- | --- |
| Title and abstract | 1 | (a) Indicate the study’s design with a commonly used term in the title or the abstract | 2 |
|  |  | (b) Provide in the abstract an informative and balanced summary of what was done and what was found | 2 |
| Introduction |  |  |  |
| Background/rationale | 2 | Explain the scientific background and rationale for the investigation being reported | 3-4 |
| Objectives | 3 | State specific objectives, including any prespecified hypotheses | 3-4 |
| Methods |  |  |  |
| Study design | 4 | Present key elements of study design early in the paper | 5 |
| Setting | 5 | Describe the setting, locations, and relevant dates, including periods of recruitment, exposure, follow-up, and data collection | 5 |
| Participants | 6 | (a) Give the eligibility criteria, and the sources and methods of case ascertainment and control selection. Give the rationale for the choice of cases and controls | 5 |
|  |  | (b) For matched studies, give matching criteria and the number of controls per case | 5 |
| Variables | 7 | Clearly define all outcomes, exposures, predictors, potential confounders, and effect modifiers. Give diagnostic criteria, if applicable | 5-6 |
| Data sources/measurement | 8* | For each variable of interest, give sources of data and details of methods of assessment (measurement). Describe comparability of assessment methods if there is more than one group | 6-7-8 |
| Bias | 9 | Describe any efforts to address potential sources of bias | 6-7-8 |
| Study size | 10 | Explain how the study size was arrived at |  |
| Quantitative variables | 11 | Explain how quantitative variables were handled in the analyses. If applicable, describe which groupings were chosen and why | 8 |
| Statistical methods | 12 | (a) Describe all statistical methods, including those used to control for confounding | 8 |
|  |  | (b) Describe any methods used to examine subgroups and interactions | 8 |
|  |  | (c) Explain how missing data were addressed | 8 |
|  |  | (d) If applicable, explain how matching of cases and controls was addressed | 8 |
|  |  | (e) Describe any sensitivity analyses | N/A |
| Results |  |  |  |
| Participants | 13* | (a) Report numbers of individuals at each stage of study—eg numbers potentially eligible, examined for eligibility, confirmed eligible, included in the study, completing follow-up, and analysed | 9 |
|  |  | (b) Give reasons for non-participation at each stage | 9 |
|  |  | (c) Consider use of a flow diagram | 9<br>Supplementary figure 1 |
| Descriptive data | 14* | (a) Give characteristics of study participants (eg demographic, clinical, social) and information on exposures and potential confounders | 9 |
|  |  | (b) Indicate number of participants with missing data for each variable of interest | N/A |
| Outcome data | 15* | Report numbers in each exposure category, or summary measures of exposure | N/A |
| Main results | 16 | (a) Give unadjusted estimates and, if applicable, confounder-adjusted estimates and their precision (eg, 95% confidence interval). Make clear which confounders were adjusted for and why they were included |  |
|  |  | (b) Report category boundaries when continuous variables were categorized |  |
|  |  | (c) If relevant, consider translating estimates of relative risk into absolute risk for a meaningful time period |  |
| Other analyses | 17 | Report other analyses done—eg analyses of subgroups and interactions, and sensitivity analyses |  |
| **Discussion** |  |  |  |
| Key results | 18 | Summarise key results with reference to study objectives |  |
| Limitations | 19 | Discuss limitations of the study, taking into account sources of potential bias or imprecision. Discuss both direction and magnitude of any potential bias |  |
| Interpretation | 20 | Give a cautious overall interpretation of results considering objectives, limitations, multiplicity of analyses, results from similar studies, and other relevant evidence |  |
| Generalisability | 21 | Discuss the generalisability (external validity) of the study results |  |
| **Other information** |  |  |  |
| Funding | 22 | Give the source of funding and the role of the funders for the present study and, if applicable, for the original study on which the present article is based |  |

\*Give information separately for cases and controls.
**Note:** An Explanation and Elaboration article discusses each checklist item and gives methodological background and published examples of transparent reporting. The STROBE checklist is best used in conjunction with this article (freely available on the Web sites of PLoS Medicine at http://www.plosmedicine.org/, Annals of Internal Medicine at http://www.annals.org/, and Epidemiology at http://www.epidem.com/). Information on the STROBE Initiative is available at http://www.strobe-statement.org.

